# Food Deserts and Food Insecurity in Indian Medical Colleges: A Mixed Methods Study Exploring Myths and Realities

**DOI:** 10.1101/2025.02.20.25322444

**Authors:** Mekha Joykutty, V. R Reshma, Anjum John

**Affiliations:** Pushpagiri Institute of Medical Sciences & Research centre, Tiruvalla, Kerala, India; Unaffiliated, freelance biostatistician

## Abstract

**Introduction:** Lack of consistent access to sufficient, nutritious food is a pressing issue globally. Resilient food systems amalgamating multiple sectors like agriculture, animal husbandry, healthcare, and education are essential to resolve food insecurity. College students, particularly in medical colleges, are vulnerable to food insecurity due to limited access to healthy food, lack of resources (time, money, and knowledge), resulting in unhealthy eating habits. This study aims to investigate the presence of food deserts within medical colleges in Kerala, analyze factors influencing student food preferences, and explore the consequences of food insecurity on their health and academic performance.

**Methods and Analysis:** A cross-sectional study will be conducted at a private medical college in Central Kerala over 8 months. Food outlets within a one-mile radius of the campus will be identified and assessed for affordability, variety, and nutritional quality. Data will be collected from medical students (medical, dental, nursing, laboratory) throughout all years using structured questionnaires, focus group discussions, and interviews with food outlet operators. Stratified random sampling will ensure representation across different academic years, and disciplines. Observations of food outlets will evaluate food variety, pricing, and hygiene standards. Quantitative data will be analyzed using descriptive and inferential statistics, while qualitative data will be thematically analyzed to gain deeper insights.

**Ethics and Dissemination:** Ethical approval has been obtained, and informed consent will be collected from participants. Data confidentiality will be strictly maintained. Findings will be disseminated through academic conferences, publications, and policy briefs using de-identified data to advocate for healthier food environments on campuses.

**Key Messages:** Food insecurity among medical college students is a relatively under-researched issue, particularly in India, with limited studies on food deserts in such settings.
This study identifies food deserts around a private medical college in Kerala and examines how limited access to nutritious food affects students’ health and academic performance.
Through a mixed-methods design, combining quantitative surveys and qualitative focus group discussions, it provides a comprehensive understanding of the food environment, including the affordability and quality of food outlets.
The findings highlight the need for policy interventions to improve food access and quality on campuses and suggest this research could guide future studies and influence public health and campus food policies.

While this study is groundbreaking for the region, its findings may be limited in generalizability to other institutions or regio

## Introduction

Food is a requirement of life, essential for survival and well-being. A well-balanced diet plays a crucial role in preventing malnutrition and keeps non communicable diseases at bay.(1)However, access to nutritious food is not universal, as it depends on availability and affordability. Food security is achieved when all people have consistent physical, social, and economic access to sufficient, safe, and nutritious food that meets their dietary needs and cultural preferences. (1,2)Conversely, food insecurity arises when there is a lack of reliable access to adequate food, compromising an active, healthy life. (3)

Resilient food systems are the solution to counter the challenges of food insecurity. A multi-dimensional approach, including agriculture, governance, health, socio-economic, and educational sectors, is required to not only counter immediate food shortages but also to establish long term food security. Among these, the education sector plays a crucial role by fostering awareness about sustainable food practices, which can contribute significantly to creating food secure campuses and empower all stakeholders, including students, to make informed choices about their nutrition. (4–6)

Food insecurity impacts school and college going students, driven by various factors. At the macro level, the affordability of nutritious food plays a major role, while meso-level factors like financial independence coupled with financial illiteracy compound the problem. Proximal factors including a lack of budgeting skills and awareness lead to food insecurity among students. Consequently, many resort to consuming unhealthy foods unbalanced diets, particularly on or near their campuses. Those who move out of parental homes are particularly vulnerable to food insecurity as they face additional challenges to accessing and affording nutritious meals. (4)

College students are in transition in various domains - navigating the shift from youth to adulthood, changes in living arrangements, body changes, and new environments. They often face barriers to accessing nutritious food, which is compounded by the pressures of balancing academic and financial responsibilities. Resource constraints including lack of time, and inadequate awareness of balanced diets force students to choose between spending on textbooks, housing near campus, transportation etc. over nutritious food. (7) This often leads to unhealthy food habits which persist in adulthood and lay the foundation for various metabolic diseases. There is a lack of access to healthy equitable food, in colleges. (8) The environments of college campuses frequently lack access to healthy, affordable, and equitable food options, effectively functioning as “food deserts” and exacerbating disparities in access to consistent, nutritious meals for young adults. (9) These changes not only result in poor dietary choices, but also affect sleep quality, academic outcomes, and overall wellbeing, leading to missed classes, and declining grades. A vicious cycle of malnutrition, micro and macronutrient imbalances, poor psychological, and physical outcomes ensue. (10) As students are the future of any country, their health is of paramount importance, and so adequate support must be provided to address basic needs like food to help them grow into healthy adults who can responsibly accept their duties as citizens of the world.

Fast food consumption is increasing in Indian campuses. (11,12) Migrations to the Middle East have altered traditional food habits, fostering unhealthy eating patterns. (13) Kerala, a southern state in India, has high health indices and top class healthcare facilities but is also fast becoming the capital of non-communicable diseases in the country. (14) The medical colleges in Kerala with their residential campuses are a home away from home for many students. (15)The duration of medical courses can extend from two years to up to 7-8 years depending on the discipline and year of studies. (16) Medical students hold the key to the health of the citizens of the country. Their health directly impacts the health of their patients. It is paramount to study the food choices of medical students and their effects on their overall performance in various aspects of their lives.(17)

This study investigates if there are food deserts within medical colleges, analyzes the various factors guiding food preferences among medical students, food insecurity in medical campuses, and gains insights into their consequences.(5) With the increasing identification of food deserts in India, this study is timely and relevant to addressing the gap in literature by increasing awareness, improving food security and health outcomes for college students.(3,18)

This study hypothesizes that medical colleges may function as food deserts, characterized by limited access to affordable and nutritious food options for students. It further posits that socio-economic factors, time constraints, and a lack of awareness significantly influence unhealthy food preferences among medical students. Additionally, the study hypothesizes that food insecurity negatively impacts students’ physical health, mental well-being, and academic performance, creating a cycle of adverse outcomes that require targeted interventions.

### Objectives

The purpose of the study is to investigate the presence of Food Deserts within medical colleges, analyze the various factors contributing to unhealthy food preferences among medical students, and assess the presence and consequences of food insecurity among students.

## Materials and Methods

The study will assess the food environment within and around a selected medical college and explore the factors influencing students’ food choices and preferences. Data will be collected using structured questionnaires, focus group discussions, interviews, and direct observations.

### Part 1: Students

#### Study Design

This is a cross-sectional design study designed to investigate food insecurity and the presence of food deserts in a medical college in Central Kerala, India.

Study setting: The study setting will be a private medical college that is accessible to the student, located in an urban area in Central Kerala, India. Because of time and resource constraints, only this one private medical college will be involved in the study at this time. Private medical colleges may have different funding sources and accessibility to food, making them an important site for exploration of the study objectives. Within the medical college, key study locations will include cafeterias and student mess halls where students commonly purchase and eat food. Eateries within the campus, and within a one-mile perimeter of the medical college (a mile radius) will be observed to assess the variety, affordability, and nutritional value of food choices available. Additionally, grocery stores accessible to medical students will be identified and evaluated to understand their contribution to the food environment.

### Sample Selection

i. Inclusion: The inclusion criteria require participants to be full-time students in medical, nursing, pharmacy, or allied health sciences programs, aged 17 years or older, proficient in English, and willing to provide informed consent.
ii. Sample frame: The sample will include medical, nursing, allied health sciences, and pharmaceutical sciences students, ensuring representation across different academic years, genders, and socio-economic backgrounds. The distribution of participants will reflect their year of study within each discipline.
iii. Rationale for the sample: Students from different academic years of various courses are planned to be included-to capture variations in food preferences, and the various academic and other challenges in different years and disciplines of study. To ensure equitable representation of students of different genders to understand gender-specific differences in food preferences and food insecurity, students from diverse socio-economic backgrounds will be included to assess the influence of financial status on food choices and access to food.
iv. Sample Size: A literature review did not identify any studies from this region that could serve as a parent study. Since the prevalence of food deserts in medical colleges in Central Kerala is not known, an estimated prevalence of food insecurity of 50% was taken, accounting for the lack of local data. The primary outcome is the prevalence of food insecurity among medical students. Using a 95% confidence interval, 5% margin of error, and Z-value of 1.96, the required sample size is estimated using the sample size formula:

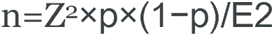

Where:

● Z = Z-value (1.96 for 95% confidence level)
● p = estimated prevalence (0.50)
● E = margin of error (0.05)

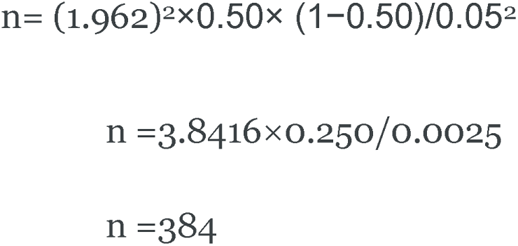

Adjusting for a 20% non-response rate, the sample size is increased to **480** students. v. Sampling Techniques: Based on literature review, a stratified random sampling suits this study. This will ensure representation across different academic years, genders, and socio-economic backgrounds. Each academic year within a college will be treated as a separate stratum. The total number of students in each stratum will be determined, and proportional sampling will be applied to select participants. Once the required proportions are established, all students within each stratum will be invited to participate, ensuring the minimum required sample size is met.

vi. Data Collection Strategies:

Questionnaire administration: Structured questionnaires will be administered to students online via Google Forms or paper-based surveys.

Questionnaire Development and Validation:

### 1.1 Content Validity

Questions will cover demographics, food access, food preferences, and perceptions of the food environment. A panel of experts from community medicine, nutrition, and behavioural sciences assessed the content validity of the questionnaire. They evaluated whether the questionnaire comprehensively covers key aspects such as food insecurity, food preferences, and related factors. Based on their feedback, necessary modifications have been made to finalize the questionnaire for use in the main study.

### 1.2 Pilot Testing

Before full-scale data collection, a pilot test has been conducted on a small group of participants to refine the questionnaire and ensure clarity. The questionnaire was evaluated for clarity, reliability, and validity during this phase. Participants in the pilot study will not be included in the final analysis. The reliability coefficient has been calculated, and necessary adjustments have been made to improve the questionnaire. A reliability coefficient above 0.7 is considered acceptable.

## 2. Actual Study

### 2.1 Questionnaire Administration (planned to be conducted soon)

The finalized structured questionnaire, developed after pilot testing, will be administered to students either online via Google Forms or through paper-based surveys. The questionnaire will cover demographic information, food availability on campus (number of food outlets, operational hours), ease of access to food (e.g., distance to outlets, food delivery options like Swiggy or Zomato), food procurement methods (direct buying vs. online ordering), variety of food (e.g., healthy vs. fast food, vegetarian vs. non-vegetarian, hot vs. packaged), and perceptions of food quality and nutritional value.

## 3. Qualitative Component: Focus Group Discussions (FGDs)

### 3.1 Conducting FGDs

If feasible, focus group discussions (FGDs) will be conducted with a diverse group of medical students to gain qualitative insights into their experiences with food availability, challenges, and suggestions for improvement. Each focus group will consist of 6 to 10 participants, ensuring a meaningful discussion while allowing all members to contribute. Discussions will be recorded (with consent), transcribed, and analysed while maintaining anonymity and confidentiality.

### 3.2 Participant Selection

Participants for FGDs will be purposively selected to ensure diversity in perspectives. Selection criteria will include year of study, gender, hostel residents vs day students, different socio-economic backgrounds. This approach will ensure a comprehensive understanding of the factors influencing food access and preferences among medical students. Discussions will focus on topics such as food options on and off campus, challenges in accessing nutritious and affordable food, and recommendations for improving the campus food environment. FGDs will be conducted in a comfortable setting to encourage open discussions, and students will lead the sessions.

#### Plan to overcome hurdles in interviews and focus group discussions: Bias Identification and Mitigation Strategies

Since participation is voluntary, students with strong opinions and willing to speak out about food insecurity may be more likely to participate. To reduce this, the study will employ stratified random sampling, ensuring representation across academic years, genders, and socio-economic backgrounds. Students may not accurately remember past food insecurity experiences. To address this, the survey will use standardized recall periods (e.g., past one month) and validated assessment tools to improve accuracy.

Participants may underreport food insecurity due to stigma. To mitigate the fact that participants might underreport food insecurity to fit into the groups, surveys will be anonymous and will be conducted in a confidential setting to encourage honesty. Students may be hesitant to divulge such information, but through establishing a rapport with them to gain a better understanding of their food choices, this hurdle may be overcome.

Given Kerala’s diverse culture, participants from different cultural, religious, ethnic, and regional backgrounds understand cultural influences on food preferences will be included. If resources permit, people working in mess halls within campus, cafeterias, and food joints within the vicinity of the college, gathering information on food availability and quality, and understanding student consumption patterns will be included in the study.

II. Food outlet operators: Part 2

Additional participants in the study will include food outlet operators-eateries and groceries- to provide contextual insights.

##### Sampling Area

A 1-mile radius will serve as the geographic boundary for identifying eligible eateries and grocery stores. A map of the area has been used to ensure that all establishments within this distance of the medical college are considered. A sampling frame of all eateries and food outlets accessible to students has been created. Depending on their numbers, a suitable sample will be studied, provided they are operational during the study period and consent to participate.

##### Gathering baseline information

Baseline information is being gathered in order to obtain the sampling frame.

##### Eateries

All types of eateries, including restaurants, fast food outlets, cafes, and street food vendors, within the 1-mile radius, through online directories, local business listings, and talking to people are identified. So far 31 eateries in the study area have been identified. Systematic sampling will be employed by selecting every nth establishment based on a predetermined interval to ensure broad coverage across the area.

##### Groceries

All grocery stores, supermarkets, and convenience stores within a 1-mile radius, again through online directories, business listings, and word of mouth are included. Seven shops, groceries, and supermarkets in the study area, all of which could be included in the study have been identified thus far.

##### Inclusion

Only eateries and grocery stores within the 1-mile radius of the medical college will be considered, and willing to provide information regarding food availability, pricing, and customer base, and who agree to onsite visits for observations.

##### Exclusion

Establishments outside the 1-mile radius or those that are not operational at the time of data collection will be excluded from the sample.

Data collection:

##### Surveys and Observations

Once the sample is selected and questionnaires finalized, the establishments will be visited to collect data on food offerings, pricing, food availability, and accessibility through surveys and observations.

##### Questionnaires

In addition to observational data, short interviews or questionnaires may be conducted with the establishment owners or managers to gather information on their food stock, supply chain, and customer base, provided they consent.

The questionnaire for eateries and grocery stores will undergo content validation by experts in nutrition, public health, and food supply management to ensure it comprehensively covers key aspects such as food availability, affordability, and supply chain operations and reliability. A pilot test will be conducted with a small sample of establishments to assess clarity, reliability, and feasibility. Necessary modifications will be made based on feedback, and a reliability coefficient above 0.7 will be considered acceptable.

##### Interviews with Food Service Providers

After questionnaires have been finalized, interviews will be conducted with owners or managers of eateries on and near campus to understand food service operations.

##### Systematic Observations

Observations of food outlets will be made to assess food variety, pricing, peak usage times, and cleanliness standards, using a checklist to ensure consistency and completeness in data collection. **Addressing Reflexivity**

Reflexivity is essential in qualitative research to acknowledge and minimize **researcher biases** that may influence data collection, interpretation, and analysis.

In this study, the following steps will be taken to ensure objective and rigorous data collection:

1. The primary researcher, a medical student, is familiar with the study environment and the challenges faced by students regarding food insecurity. This familiarity can help build trust with participants, but it may also introduce preconceptions or biases. To minimize this, the researchers will use open-ended, non-leading questions in interviews and focus group discussions. Keeping a journal to document personal assumptions, or biases that occur during data collection would be encouraged. With experienced researchers, we will try to conduct peer debriefing sessions to critically reflect interpretations and coding. To get a complete picture of the situation of food security around the students, the study will employ multiple data sources, including individual interviews, Focus Group Discussions, observations of food outlets, and interviews with food service providers. Cross-checking information from different sources, to help reduce researcher biases and maintain accuracy. Transcribed interviews and focus group discussions will be coded independently by two researchers, and discrepancies in coding will be resolved through discussion. This approach ensures interpretative consistency and minimizes subjective bias in thematic analysis.

### Outcomes, Predictors, Confounders, and Effect modifiers

Given the lack of literature on this topic, the proposed outcomes, predictors, confounders, and effect modifiers are based on theoretical assumptions and potential factors identified through existing literature on food preferences, food insecurity, and socio-economic influences in other settings. These elements will be explored in the context of this study, and their relevance will be assessed as data collected. The study will not include any experimental interventions.

Instead, it will measure as outcomes:

1. **Food Preferences:** Types of food preferred by students (e.g., vegetarian vs. non-vegetarian, processed vs. whole foods, cultural food preferences), Frequency and variety of food consumed, Nutritional quality of food choices.
2. **Food Insecurity:** Defined as limited or uncertain access to adequate food. **Severity of food insecurity**: Mild, moderate, or severe levels **Access to food:** Number of meals skipped, reliance on food assistance, food budgeting, etc. Food insecurity levels using a validated food security questionnaire created for the study(attached)
3. **Impact on Health and Well-being:** Measures of overall well-being (e.g., stress levels, mental health, self-reported physical health), Academic performance (e.g., attendance, focus during classes and test marks).

Predictors: These are the factors that are hypothesized to influence the outcomes:

1. **Gender:** may exhibit different food preferences and vulnerabilities to food insecurity.
2. **Socio-economic Status:** Family income, parental education, status, or scholarship status.
3. **Cultural Background:** Cultural beliefs and religious practices regarding food, which may influence preferences and food-related behaviors.
4. **Accessibility of Food:** Proximity to affordable eateries and grocery stores, availability of healthy food options in the vicinity.
5. **Academic Pressure:** Time constraints or stress related to academics may influence food choices (e.g., reliance on fast food, skipping meals).
6. **Student’s Level of Food Literacy:** Knowledge about nutrition, accessibility to cooking facilities, and the ability to make informed food choices.

#### Confounders

These are variables that might influence both the predictor and outcome, leading to potential bias-these are again hypothesized as:

1. **Age:** Younger students may have different food choices and vulnerabilities compared to older students.
2. **Living Arrangements:** Whether students live in hostels, with families, or independently can affect their food choices and access to food.
3. **Health Status:** Pre-existing health conditions (e.g., diabetes, obesity) may influence food preferences and food insecurity.
4. **Location:** The proximity of the student’s residence to available resources may affect food access. Transportation access may again affect food procurement.

#### Effect Modifiers

These are factors that are hypothesized to modify the strength or direction of the relationship between the predictor and the outcome:

1. **Gender:** The relationship between socio-economic status and food insecurity may vary by gender due to differing societal roles and expectations.
2. **Cultural Factors:** Cultural preferences and religious practices may amplify or dampen the effects of socio-economic factors on food choices and food insecurity.
3. **Social Support:** Availability of family or peer group shared meals may influence the effect of food insecurity on well-being.
4. **Academic Year:** The year in which the student is enrolled (e.g., first-year students may experience different stress and food insecurity than final-year students) and the academic discipline of study.

### Timeline of the study

The duration of the study will be about 8 months

The timeline of the study will be.

#### Months 1-2

Getting administrative approvals from the medical college authorities,

and identifying food joints, supermarkets, and other food places within the vicinity of the medical college.

#### Month 3-6

Conduct surveys, in-depth interviews, focus groups and follow-up surveys.

#### Month 7

Data analysis and interpretation.

#### Month 8

Report writing and dissemination of findings.

These activities will help delineate the food environment surrounding the selected medical college and help us understand whether there are food deserts in and around the medical college and if there is food insecurity faced by medical students at that college.

### Patient and Public Involvement

Patients or the public WERE NOT involved in the design, or conduct, or reporting, or dissemination plans of our research.

Patients and the public were not involved in any way in this protocol. Eatery and grocery owners will be participants in the research.

### Data Analysis plan

Quantitative data will be analyzed using SPSS software: Descriptive statistics will summarize demographic and food security variables. Chi-square tests to explore associations between categorical variables. Logistic regression to identify predictors of food insecurity. The subgroup analyses to examine variations in food insecurity by gender, academic year, and socio-economic background.

Qualitative data from focus group discussions and interviews will be transcribed verbatim and analyzed thematically using NVivo software. Emergent themes will be identified to provide deeper insights into the factors contributing to food insecurity and its consequences.

By integrating quantitative and qualitative data, this study aims to provide a comprehensive understanding of food insecurity and its determinants in medical college settings, informing policies to promote healthier food environments.

### Operational Definitions

1. Food insecurity: Is when people do not have enough to eat and do not know where their next meal will come from. Food insecurity will be classified as: high food security (lowest food insecurity, minimal), marginal food security (medium food insecurity, stressed), low food security (high food insecurity, crisis), very low food security (high food insecurity, emergency).
2. Food insecurity domains: Availability of food, Access to food, Utilization of food, and Stability of food.

a. Food availability: Physical availability of food
b. Food access: Economic and physical access to food
c. Food utilization: This includes all stages of food handling and preparation from production until it is eaten. It includes procurement of food through processing of food, packing and storage, food transport, food preparation, the diversity of the diet, and how food is distributed among the consumers of prepared food. After that it includes good biological utilization of the food once consumed, which determines the nutritional status of the person.
d. Food Stability: The availability, access, and utilization of food over a prolonged duration of time.
3. Food desert: An area where people have limited access to wholesome, healthy food of variety, at affordable rates. The shops have limited shelves, so only foods so they can store only packaged or processed foods with long shelf lives. Theoretically, living in a food desert is associated with a higher risk of poor nutrition and makes people prone to lifestyle diseases like obesity, diabetes, heart disease, or leads to metabolic diseases of various types.
4. Food Swamps: retail shops that dispense unhealthy foods like fast foods, more than healthier alternatives.
5. Eatery: A building where people go to eat.
6. Grocery store: A food retail store - where different types of food items are sold.
7. Medical Student: A person enrolled in a medical course that gives them a degree in medicine. For this research, nursing, laboratory, and pharmacy students will be considered as medical students.

### Ethical Issues

Data will only be collected from students who have willingly given their consent. The data which collected from them will be highly confidential and will not lead to any sort of discrimination. Data collected from eateries and food outlets will also be kept confidential and private. The study does not aim to question the preferences of the students by keeping in mind their individual rights but rather the purpose is to find out about the food security status of a medical college in Central Kerala.

### Implications

Food insecurity is a growing problem around the globe affecting a variety of people belonging to different classes. College students are one such group which are affected by food insecurity. The impacts and consequences of food insecurity among college students are not just restricted to their health but it extends to other domains. Some of the negative impacts of food insecurity faced by college students are effects on academic performance, physical health, and mental health. Diets are influenced by several factors which include income, food prices, food preferences, beliefs and traditions, or geographical and environmental factors. Hence advocating for a healthy food environment is crucial for maintaining healthy individuals in society. To unveil whether food deserts exist within Indian medical colleges is the aim of the study, which will enable the college administrators to understand the current food security status in medical students who are the future of medicine.

### Work done so far

M.J., a medical student, participated in writing parts of the proposal, doing literature review, gathering data the sampling frame for the students and eateries, pilot testing of questionnaires, reviewing protocol before submission.

A.J. is the guide of the student, who conceived the idea, supervised the protocol writing, literature review, ethical approval process, funding submission to ICMR, and the current publication process to the journal.

RVR is the consultant biostatistician on the project, who reviewed the protocol and helped design the sampling frame, and the sampling methods for the protocol.

## Data Availability

Not applicable

